# Effectiveness of DiabeText, a mHealth intervention to support diabetes self-management: randomized controlled trial in primary care

**DOI:** 10.1101/2024.02.28.24303489

**Authors:** Rocío Zamanillo-Campos, Maria Antonia Fiol-Deroque, Maria Jesús Serrano-Ripoll, Joan Llobera-Canaves, Joana María Taltavull-Aparicio, Alfonso Leiva-Rus, Joana Ripoll-Amengual, Escarlata Angullo-Martínez, Isabel Maria Socias-Buades, Luis Masmiquel, Jadwiga Konieczna, María Zaforteza-Dezcallar, Maria Asunción Boronat-Moreiro, Sofía Mira-Martínez, Elena Gervilla-García, Ignacio Ricci-Cabello

## Abstract

**Background:** Complications arising from uncontrolled Type 2 Diabetes (T2D) poses a significant burden on individuals’ well-being and healthcare resources. Digital interventions may play a key role in mitigating such complications by supporting patients to adequately self-manage their condition.

**Aim:** To assess the impact of DiabeText, a new theory-based, patient-centered, mobile health intervention integrated with electronic health records to send tailored short text messages to support T2D self-management.

**Design and setting:** Pragmatic, phase III, 12-month, two-arm randomized clinical trial with T2D primary care patients in Spain.

**Method:** 742 participants with suboptimal glycemic control (HbA1c>7.5) were randomly allocated to a control (usual care) or intervention (DiabeText) group. The DiabeText group received, in addition to usual care, 165 messages focused on healthy lifestyle and medication adherence. Primary outcome: glycated hemoglobin (HbA1c). Secondary outcomes: medication possession ratio, quality of life (EQ-5D-5L), diabetes self-efficacy (DSES); and self-reported adherence to medication, Mediterranean diet (MEDAS-14), and physical activity (IPAQ).

**Results:** At 12 months follow-up, no statistically significant differences in mean HbA1c were observed between the intervention (7.5 [95%CI 6.7 to 8.2]) and control groups (7.4 [6.7 to 8.3]). In comparison with the control group, the DiabeText group showed significant (p<0.05) improvements in self-reported medication adherence (OR=1.4; 95%CI: 1.0 to 1.9), DSES (Cohen’s d=0.4), and EQ5D-5L (Cohen’s d=0.2) scores; but not in the rest of secondary outcomes.

**Conclusion:** DiabeText successfully improved quality of life, diabetes self-management, and self-reported medication adherence in primary care patients with T2D. Further research is needed to enhance its effects on physiological outcomes.

## Introduction

Type 2 diabetes (T2D) is a chronic condition that is among the top causes of premature death. People with T2D are at high risk of developing serious complications (e.g., blindness, lower-limb amputations, kidney disease, and cardiovascular disease), which reduce their quality of life and life expectancy. The cornerstone of T2D management is promoting a healthy lifestyle and taking glucose lowering drugs; but adherence to treatment plans is the truly bottleneck for achieving good glycemic control in patients with T2D. Hence, there is an urgent need to find highly implementable and effective interventions to promote adherence to treatments in this population.

Over the last decade, advances in technology and connectivity have led to a boom of mobile health (mHealth) interventions, as they constitute a low-cost and highly scalable approach to support behavior changes in terms of lifestyle ^1–4^ and medication taking ^5^. However, as highlighted by the European Association for the Study of Diabetes (EASD) and the American Diabetes Association (ADA), while the available studies of m-Health interventions show promise for promoting healthy behaviour and managing complex diseases, such as T2D, they are very limited in both quantity and quality. According to the most recent systematic reviews and meta-analysis, there is an urgent need for large, adequately powered RCTs, to examine longer term efficacy before promoting the adoption and wide-spread dissemination of such interventions.

The aim of this study was to assess the effectiveness of DiabeText, a new theory-based, patient-centered, mobile health intervention integrated with electronic health records to send tailored short text messages to support T2D.

## Methods

### Design

Pragmatic, phase III, 12-month, two-arm (1:1 allocation ratio), randomized clinical trial with T2D primary care patients in Spain. We adhered to the CONSORT 2010 Statement updated guidelines for reporting parallel group randomized trials. A full study protocol is available elsewhere^6^.

### Study participants

We included patients with T2D registered in the public health service of the Balearic Islands; aged >18 years; with HbA1c levels >7.5% recorded during the previous 6 months; with at least one prescription of a non-insulin antidiabetic drug, and; able to receive, read and understand SMS in Spanish through a personal mobile phone. We excluded people with severe mental conditions or participating in other research studies.

### The DiabeText Intervention

Participants allocated to the intervention (DiabeText) group were sent 167 SMS during the 12 months follow-up period. These messages were distributed as follows: 5 SMS per week during 4 months, 3 SMS per week the next 4 months and 2 SMS per week the last 4 months. The DiabeText intervention was developed based on the MRC guidelines for the development of complex interventions. The development process involved systematic reviews, qualitative studies with patients and primary care providers, and a phase two feasibility trial. The messages content (developed by a multidisciplinary team, including patients) was related to diabetes management and the use of medicine. Participants also received extra messages with the latest blood test results, changes in body weight and reminders for medical appointments and pharmacy medication pickup. The complete template for DiabeText intervention description and replication checklist^7^ is available as **Supplementary Material 1**.

### Sample size

Sample size was estimated as 740 participants (370 per group) to detect changes in HbA1c between groups of 4 mmol/mol (0.4%) based on a standard deviation of 15mmol/mol (1.5%). This estimate includes a 20% loss to follow-up at 90% power and p=0.05.

### Recruitment and baseline data collection

With support from an information specialist (see acknowledgements), we extracted a list of patients potentially meeting the eligibility criteria based on data recorded in e-SIAP, the primary care electronic health record system. We sent these potential participants an SMS to their mobile phones inviting them to participate in the study. Subsequently, trained research assistants phoned them to seek confirmation of eligibility criteria, and to record informed consent when appropriate. Baseline data was then collected through semi-structured phone interviews.

### Randomization and blinding

Randomization was performed using the Spanish adaptation of the free OxMaR software^8,9^. We applied a non-deterministic minimization algorithm to ensure that the intervention groups were balanced on important baseline prognostic factors (age and sex). Patients were informed on their allocation to the control (receiving usual care) or the intervention (usual care + DiabeText) group only after all baseline data had been collected. Data collectors and analysts were blinded.

### Outcomes

The primary outcome was HbA1c (%) at 12-month follow-up (post-intervention). Secondary outcomes included medication possession ratio (MPR), self-reported adherence to diabetes medication^10^, health-related quality of life (EQ-5D-5L)^11,12^ ; diabetes self-efficacy (DSES) ^13^; adherence to Mediterranean diet (MEDAS-14)^14^, and physical activity (IPAQ) ^15^. We used ad hoc questionnaires and questions to measure users’ satisfaction, perceived utility, perceived ease access and potential harms related to the intervention. A detailed description on outcomes measures and methods for data collection are available in **Supplementary Material 2**.

### Statistical analysis

Mean and standard deviations (sd) or median and interquartile range (IR) for continuous variables, and frequencies and percentages (%) for categorical variables were calculated for descriptive analysis of the participants’ socio-demographic, lifestyle and health-related characteristics. The effect of the intervention was evaluated using general linear models or logistic regression as appropriate, adjusted for corresponding baseline values. We checked the robustness of our findings to non-normality using non-parametric bootstrapped test ^16^. Main analyses were performed using multiple imputation chained equations (MICE) models with 10 iterations and bootstrap inference^17^. Results were unchanged when using the evaluable population (**Supplementary Material 3**). The adjusted mean difference between groups is presented along with its associated 95% CI and p-value. Cohen’s D was calculated to estimate effect sizes for significant results. All the analyses were carried out on the basis of intention-to-treat. Subgroup analyses (sex, age group (<65 vs ≥65 years old) and medication taking at baseline (non-adherent vs adherent) based on MPR (<80% vs ≥80%) and self-reported) were performed when statistically significant interactions were found. All analyses were carried out in Stata 15 (StataCorp) and an α of 5% throughout was used as a threshold for significance.

## Results

### Recruitment and baseline characteristics

Between 6^th^ October and 18^th^ November 2021 we sent SMS invitations to 2,218 potentially eligible participants (Figure 1). Among the 1,591 patients who were successfully contacted via telephone, and who confirmed meeting the eligibility criteria, 742 agreed to participate (recruitment rate = 47%). At twelve months follow-up, 68 patients declined to participate in the final phone interview or were not reachable over the phone (withdrawal rate = 9.2%), whereas 674 patients (340 in the control and 334 in the intervention group) completed the post-intervention assessment. The 95.2% participants in the intervention group received more than 90% of the intervention (>165 messages in one year).

**Figure 1.**
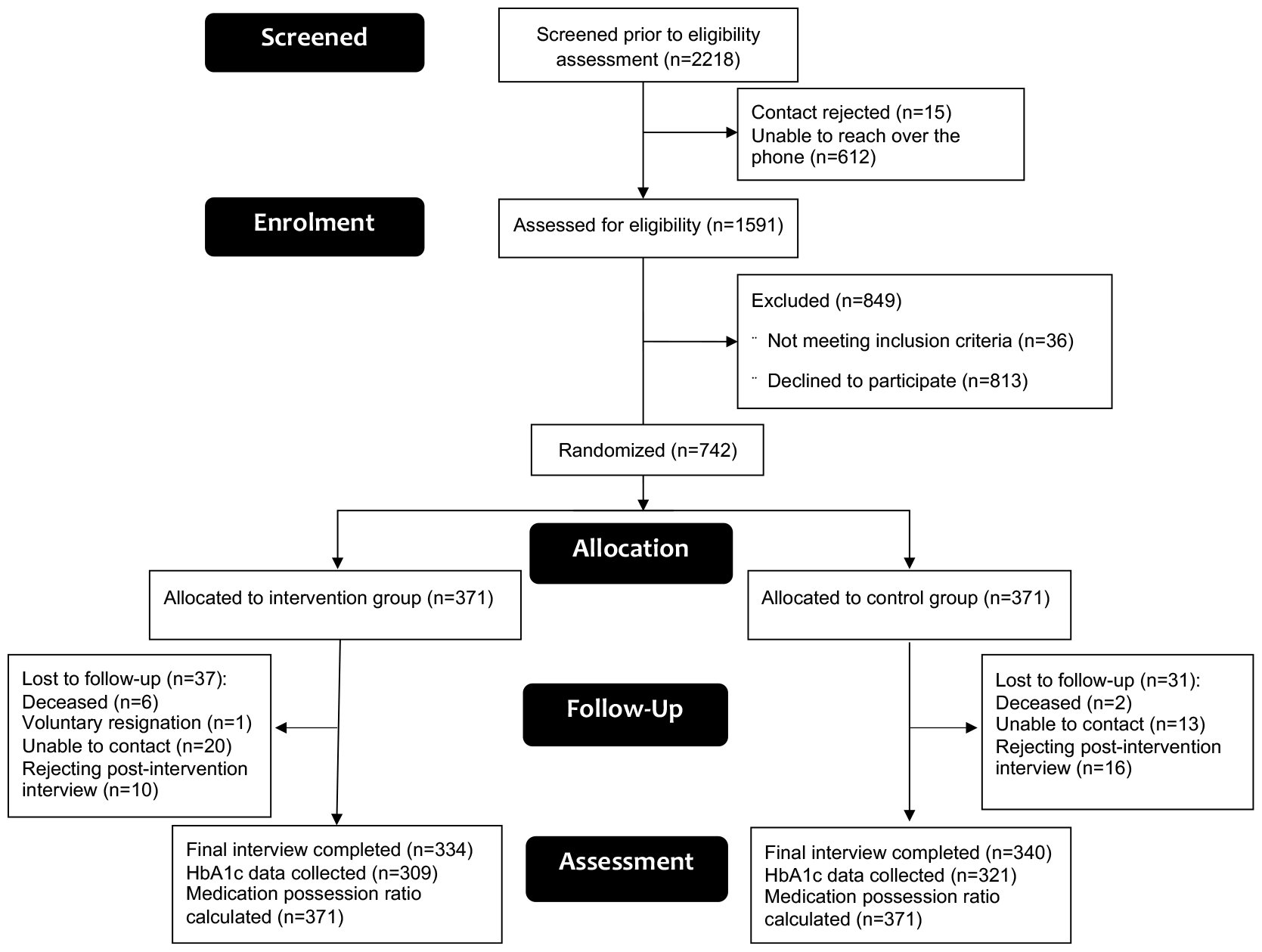
Consort flowchart of the inclusion of patients in the DiabeText trial.

The baseline sociodemographic and clinical characteristics of the 742 participants are shown in Table 1. About 41.6% were women (309/742), with a mean (sd) age of 65 (10) years and 10 (5) years since the diagnosis of T2D. More than a half of them (N=428, 57.7%) had obesity (BMI>30), around two-thirds (N=513, 69.1%) had hypertension, and one quarter were current smokers (N=178, 24%). The mean (sd) number of active antidiabetic prescriptions was 1.72 (0.71). Median (IQR) HbA1c was 8.1% (7.7-8.8%). We observed no relevant differences in baseline sociodemographic and clinical characteristics between the intervention and control groups; and between participants with complete or not complete follow-up data (data not shown).

**Table 1.** Baseline sociodemographic and clinical characteristics of the study participants.

|  | Overall<br>(n=742) | Control group<br>(n=371) | Intervention group<br>(n=371) |
| --- | --- | --- | --- |
| Sociodemographic characteristics |  |  |  |
| Female, n (%) | 309 (42) | 154 (42) | 155 (42) |
| Age (years), mean (sd) | 65 (10) | 65 (10) | 65 (10) |
| Current smoker, n (%) | 178 (24) | 84 (24) | 94 (27) |
| Reduced mobility, n (%) | 24 (3.2) | 8 (2.2) | 16 (4.3) |
| Use of Internet in the mobile phone, n (%) | 537 (72) | 269 (73) | 268 (72) |
| Clinical characteristics |  |  |  |
| Diabetes duration (years), mean (sd) | 10.3 (5.2) | 10.3 (5.2) | 10.4 (5.3) |
| Caregiver, n (%) | 51 (6.9) | 29 (7.8) | 22 (5.9) |
| HbA1c (%), median (IQR) | 8.1 (7.7-8.8) | 8.0 (7.6-8.8) | 8.1 (7.7-8.7) |
| Medication Possession Ratio (%), median (IQR) | 95.6 (82.4-99.3) | 96.2 (83.1-99.3) | 95.1 (81.8-99.3) |
| Number of antidiabetic active substances, mean (sd) | 2.3 (0.9) | 2.2 (0.8) | 2.4 (0.9) |
| Number of antidiabetic prescriptions, mean (sd) | 1.72 (0.71) | 1.67 (0.69) | 1.77 (0.73) |
| Number of overall prescriptions, mean (sd) | 7.2 (3.8) | 7.0 (3.6) | 7.4 (4.0) |
| Hypertension, n (%) | 513 (69) | 250 (67) | 263 (71) |
| Obesity (BMI>30) n (%) | 428 (57.7) | 219 (59.0) | 209 (56.3) |
| Depression, n (%) | 164 (22.1) | 90 (24.3) | 74 (19.9) |
| Diabetic foot, n (%) | 64 (8.6) | 33 (8.9) | 31 (8.4) |
| Chronic kidney disease, n (%) | 87 (11.7) | 51 (13.8) | 36 (9.7) |
| Diabetic retinopathy, n (%) | 20 (2.7) | 9 (2.4) | 11 (3.0) |
| Diabetic neuropathy, n (%) | 20 (2.7) | 7 (1.9) | 13 (3.5) |

BMI, body mass index; IQR, interquartile range; sd, standard deviation; HbA1c, glycated haemoglobin

### Glycemic control

Follow-up HbA1c data was successfully collected from 630 of the 742 participants in the study (84.9%). At baseline, no significant differences were observed between the intervention (8.1 (7.7-8.7)) and control group (8.0 (7.6-8.8)). Over the 12-month period, there was a similar reduction in HbA1c values in both groups (7.4% and 7.5% in the control and intervention group, respectively), with no significant differences between groups observed (Table 2).

**Table 2.** Association between the DiabeText intervention and glycemic control, adherence to medication treatment, diabetes self-efficacy and quality of life.

|  | Baseline |  | 12 months follow-up |  | Association coefficient estimates for participants in the intervention group compared to controls |  | Effect size |
| --- | --- | --- | --- | --- | --- | --- | --- |
|  | Control (n=371) | Intervention (n=371) | Control (n=340) | Intervention (n=334) | Beta (95%CI) | p-value<sup>1</sup> | Cohen's D (95%CI) |
| Glycemic control (HbA1c (%)), median (IQR) | 8.0 (7.6 to 8.8) | 8.1 (7.7 to 8.7) | 7.4 (6.7 to 8.3)<sup>2</sup> | 7.5 (6.7 to 8.2)<sup>2</sup> | -0.028 (-0.200 to 0.145) | 0.750 | N/A |
| Adherence to medication treatment (MPR (%)), median (IQR) | 96.2 (83.1 to 99.3) | 95.1 (81.8 to 99.3) | 91.0 (73.6 to 97.8)<sup>3</sup> | 90.4 (76.2 to 97.7)<sup>3</sup> | -0.266 (-2.077 to 1.542) | 0.773 | N/A |
| Diabetes self-efficacy scale (DSES), median (IQR) | 6.9 (5.9 to 7.9) | 6.9 (5.8 to 8.0) | 8.0 (6.6 to 8.9) | 8.6 (7.4 to 9.3) | 0.544 (0.314 to 0.773) | **<0.001\*** | 0.35 (0.20 to 0.50) |
| Quality of life (EQ-5D index<sup>18</sup>), median (IR) | 0.93 (0.89 to 1) | 0.93 (0.88 to 1) | 0.97 (0.89 to 1) | 1 (0.92 to 1) | 0.015 (0.003 to 0.028) | **0.012\*** | 0.18 (0.04 to 0.32) |

Analyses were performed using linear mixed-effects models adjusted for baseline values with MI BOOT estimations.
HbA1c, glycated haemoglobin; IQR, interquartile range; CI, 95% coefficient interval; N/A, not applicable
<sup>1</sup> p values and 95%CI based on percentiles using non-parametric Bootstrap
<sup>2</sup> post-intervention HbA1c available for 321 patients in the control group and 309 in the intervention group
<sup>3</sup> post-intervention MPR available for 371 patients in each group respectively, so no imputations were necessary
\* p<0.05

### Medication possession rate and self-reported medication adherence

Follow-up MPR data was successfully collected for all the study participants (n=742). At baseline, no significant differences were observed between the intervention and control groups. Over the 12-month period, there was a reduction in MPR values in both groups: from median 96.2% and 95.1% at baseline, to 91% and 90.4% at 12 months in the control and intervention group, respectively. No significant differences between groups were observed at the 12 months follow-up (Table 2). Similarly, no significant differences were observed when after dichotomizing MPR to compare adherent (MPR ≥ 80%) vs. non-adherent (MPR < 80%).

Regarding self-reported adherence to antidiabetic medication, there was a statistically significant improvement in the intervention group compared to control group (OR=1.41; 95%CI 1.03 to 1.92; p=0.029). A significant interaction with sex was found, showing that between-group differences were greater (p for interaction = 0.037) in men (OR=1.87, 95% CI 1.24 to 2.8, p=0.002) than in women (OR=0.95, 95% CI 0.58 to 1.55, p=0.838) (Table 3).

**Table 3.** Association between the DiabeText intervention and medication possession ratio, self-reported adherence to antidiabetic medication, adherence to Mediterranean diet and adherence to physical activity.

|  | Baseline |  | 12 months follow-up |  | Association odds for participants in the intervention group compared to controls |  |
| --- | --- | --- | --- | --- | --- | --- |
|  | Control (n=371) | Intervention (n=371) | Control (n=340) | Intervention (n=334) | OR (95% CI)<sup>1</sup> | p-value<sup>2</sup> |
| **Medication Possession Ratio<sup>3</sup>, n (%)** |  |  |  |  |  |  |
| Non-adherent (MPR < 80%) | 84 (22.6%) | 89 (24%) | 113 (30.5%) | 116 (31.3%) | 1 |  |
| Adherent (MPR ≥ 80%) | 287 (77.4%) | 282 (76%) | 258 (69.5%) | 255 (68.7%) | 0.987 (0.696 to 1.400) | 0.943 |
| **Self-reported adherence to antidiabetic medication, n (%)** |  |  |  |  |  |  |
| Non-adherent | 177 (47.7%) | 166 (44.7%) | 143 (42.1%) | 114 (34.1%) | 1 |  |
| Adherent | 194 (52.3%) | 205 (55.3%) | 197 (57.9%) | 220 (65.9%) | 1.409 (1.034 to 1.920) | **0.029\*** |
| **Adherence to Mediterranean diet, n (%)** |  |  |  |  |  |  |
| Non-adherent | 90 (24.3%) | 100 (27.0%) | 171 (50.3%) | 163 (48.8%) | 1 |  |
| Adherent | 281 (75.7%) | 271 (73.0%) | 169 (49.7%) | 171 (51.2%) | 1.11 (0.829 to 1.486) | 0.481 |
| **Adherence to physical activity, n (%)** |  |  |  |  |  |  |
| Non-adherent (low level) | 126 (34.0%) | 131 (35.3%) | 146 (43%) | 163 (48.8%) | 1 |  |
| Adherent (moderate or high level) | 245 (66.0%) | 240 (64.7%) | 194 (57%) | 171 (51.2%) | 0.824 (0.600 to 1.132) | 0.234 |

Analyses were performed using logistic regression adjusted for baseline values with MI BOOT estimations.
<sup>1</sup> data represents Odds Ratio (OR) with 95% confidence intervals in parentheses (CI)
<sup>2</sup> p values and 95%CI based on percentiles using non-parametric Bootstrap
<sup>3</sup> post-intervention MPR available for 371 patients in each group respectively, so no imputations were necessary
\*Statistically significance stated at p<0.05

### Lifestyle behavior, diabetes self-efficacy and health-related quality of life

Follow-up data on lifestyle behavior was successfully collected for 674 participants. At baseline, no significant differences were observed between the intervention and control groups in terms of the proportion of participants with low adherence to the Mediterranean diet (27% and 24%) and with low levels of physical activity (35% vs. 34%), respectively. Over the 12-month period, no significant differences were observed in adherence to the Mediterranean diet (OR = 1.11; 95% CI 0.83 to 1.49) and physical activity (OR = 0.83; 95% CI 0.70 to 1.13) (Table 3).

At baseline we observed no significant differences in median (IQR) DSES scores between the intervention (6.9 (5.8-8.0)) and control group (6.9 (5.9-7.9)). At 12 months follow-up DSES scores were significantly (p<0.001) higher in the intervention (8.63 [7.38-9.25]) than in the control group (8.0 [6.63-8.88]). A Cohen’s d of 0.35 (95% CI 0.20 to 0.50) indicated that the effect size was small (Table 2).

At baseline we observed no significant differences in median (IQR) EQ-5D scores between the intervention (0.93 (0.88 to 1.00)) and control group (0.93 (0.89 to 1.00)). At 12 months follow-up EQ-5D scores were significantly (p<0.05) higher in the intervention (1.00 [0.92 to 1.00]) than in the control group (0.97 [0.89 to 1.00]). A Cohen’s d of 0.18 (95% CI 0.20 to 0.50) indicated a small effect size in health-related quality of life (Table 2).

### Participants’ satisfaction with the intervention

Participants in the intervention group perceived DiabeText as a useful resource to help them manage their diabetes (mean (sd) perceived utility score 8.1/10 points (2.47)). The ease of access to the messages was high, with a mean (sd) score of 8.8/10 points (2.09). In general, participants were satisfied with the DiabeText intervention (mean satisfaction score 8.1/10 (2.37)). Two participants perceived that that the messages were intrusive and produced anxiety. The remaining 332 participants did not report any harm or adverse effects associated with DiabeText.

## Discussion

### Summary

This trial shows that after 12 months DiabeText, a personalized mHealth intervention to support T2D self-management, did not produce a significant effect on glycaemic control, medication possession ratio and lifestyle behaviour. However, DiabeText was associated with improvements in adherence to antidiabetic medication, diabetes self-efficacy, and health-related quality of life.

### Strengths and limitations

This randomized controlled trial (RCT) includes a large and representative sample of people with T2D, which allows to generalize the results. Moreover, internal validity is warranted by the randomization and blinding process. Also, the study used validated and reliable tools for data collection; objective measures were collected, such as HbA1c and MPR and; the intervention was designed and developed following a standardized procedure. In addition, the intervention was based on official guidelines and the latest scientific evidence regarding diabetes treatment and monitoring.

In terms of limitations, first, instead of ordering laboratory tests to measure HbA1c, we followed a pragmatic approach, extracting this data from electronic clinical records when available. We only ordered blood tests when data were not available. This fact could have delayed part of the measurements until the end of the follow-up. However, as indicated in supplementary material 2, we predefined a maximum of 3 months before and after the end of the intervention to reduce the time range and avoid dilution of the possible effects of the intervention. It is important to note here that HbA1c measurements represent blood glucose levels from the previous 3 months. Second, in relation to the use of MPR as an outcome variable, is worth noting that medication possession does not necessarily imply medication taking. However, no gold standard exists and using MPR is more reliable ^19^.

### Comparison with existing literature

A recent meta-analysis of nine trials evaluating the effect of SMS interventions vs usual care on glycaemic control ^20^ showed a pooled effect on HbA1c of -0.37% [-0.57 to -0.17]. This reduction is relatively small, and three of these studies did not observe statistically significant reductions. Similar results were previously reported by Haider *et al*. (−0.38%[-0.53 to -0.23])^21^. Taken together, these findings indicate that some, but not all SMS interventions to support diabetes self-management are effective in reducing HbA1c. More research is needed to better understand what factors predict stronger benefits in glycaemic control. In addition to the above mentioned, Moschonis *et al*. observed a clinically meaningful reduction (>0.5%) in HbA1c in the intervention group for 9 out of 20 studies and in the control group for 5 out of 20 studies evaluating SMS interventions ^20^. This is exactly what we observed in our study: a -0.6% reduction in both groups at 12 months follow-up. This suggests a strong Hawthorn effect, possibly exacerbated by the fact that the recruitment of the participants took place only several months after the covid-19 pandemic lockdown– period during which the proportion of poorly controlled HbA1c substantially increased.

Available evidence shows that telehealth interventions are an effective method to increase medication adherence in patients with T2D. In regard to effectiveness of SMS on medication adherence, a recent meta-analysis^5^, which included nine studies and 1,121 participants, showed a pooled effect size of 0.36 (p=0.001). In our study, which included 742 participants, the intervention effect on self-reported medication adherence was very similar (OR=1.41; p=0.03). However, when adherence was estimated in terms of MPR, we did not observe significant differences between groups while Vervloet *et al* found them^22,23^. It is possible that including people with good adherence (MPR>82%) leaves little room for improvement. It also reinforces the idea that dispensing the medication from the pharmacy does not necessarily imply taking such medication as instructed by the prescribers.

Regarding the active ingredients of SMS interventions we reviewed ^5,20,21^, DiabeText exhibited similarities in terms of intervention duration (ranging from 3 to 24 months) and message frequency (ranging from daily to twice a week). Moreover, the content of the messages closely resembled that of DiabeText, with only two exceptions where the content focused on medication adjustments in response to blood glucose levels. Six studies incorporated multiple components (e.g., booklet, web-based portal, mobile application, phone calls, in-person training, or picture messages). However, the sample sizes of prior studies were consistently less than half the number of participants included in our study, except for the INDICA study, which enrolled 2,334 participants^24^. Considering the heterogeneity across studies and the limited number of individual studies reporting significant intergroup differences, the DiabeText study is poised to enhance the robustness of emerging findings.

We observed that DiabeText was associated with improvements in diabetes self-efficacy and health-related quality of life. These outcomes are measured less frequently, with different instruments and usually as secondary outcomes. Consequently, current evidence is still scarce and inconsistent, with some studies observing significant improvements ^25–28^ while others did not^29,30^.

Similarly, dietary and physical activity estimates are scarcely reported and measured with incomparable instruments^26,28^. Although we observed a trend to improvement in our previous feasibility trial^31^ it was not the case in the current effectiveness trial as observed in similar studies^28,32,33^. A recent systematic review showed inconsistent results of individual studies regarding the effectiveness of mobile text messaging on physical activity in people with T2D, highlighting the need for more high-quality primary studies in this area.

### Implications for research and clinical practice

Considering that SMS interventions have been evaluated as effective for improving medication taking and glucose control by recent meta-analysis and that DiabeText improve quality of life, diabetes self-efficacy and self-reported adherence to drug treatments, clinicians and other stakeholders should familiarise with SMS interventions and consider the prescription of DiabeText to support caring of people with T2D.

In the meantime, future studies are needed to determine how to maximize the benefits of mHealth interventions, especially on physiological outcomes such as glycaemic control. In that sense, a recent meta-analysis provided evidence that mHealth is likely to be beneficial for diabetes patients when the right behaviour change techniques are applied to realize the full advantage of the intervention. Further investigation of the role of theory in the design of mHealth interventions is warranted. More work is also needed to investigate how individual demographic, socioeconomic, behavioural and clinical characteristics contribute to patient engagement and the efficacy of mHealth tools for diabetes.

## Data Availability

All data produced in the present study are available upon reasonable request to the authors

## Acknowledgments

We are grateful to Professor Andrew Farmer for his ideas and insight in the initial development of this work. The authors thank the central services from the Balearic Island public health services for they support and help with their software to send text messages to patients. We thank Pau Pericàs, from the PRISIB (health data platform from the IdISBa health research institute), for his support programming the randomization software and extraction of participants health data to personalise the messages.

