## Supplementary material 1 for "Effectiveness of DiabeText, a mHealth intervention to support diabetes self-management: randomized controlled trial in primary care"

### Supplementary material 1. Detailed description of the DiabeText intervention (TiDieR checklist).

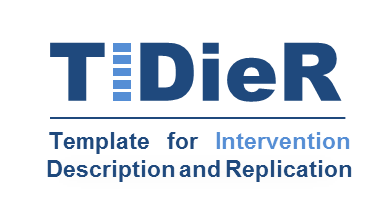
**The TIDieR (Template for Intervention Description and Replication) Checklist*:**

| Item | | Description |
| --- | --- | --- |
| 1. | **BRIEF NAME**  Provide the name or a phrase that describes the intervention. | DiabeText is a mobile health Intervention to Support Diabetes Medication taking in adults with Type 2 Diabetes receiving antidiabetic treatment |
| 2. | **WHY**  Describe any rationale, theory, or goal of the elements essential to the intervention. | To promote diabetes self-management including medication taking and healthy lifestyles in people with type 2 diabetes mellitus |
| 3. | **WHAT**  Materials: Describe any physical or informational materials used in the intervention, including those provided to participants or used in intervention delivery or in training of intervention providers. Provide information on where the materials can be accessed (e.g. online appendix, URL). | Tailored short text messages (SMSs) sent to the mobile phones of adults with type 2 diabetes. Participants are grouped in three different profiles based on their lifestyle characteristics coming from the 14 items short screener for assessing Mediterranean Diet Adherence among adults (MEDAS-14) and que International Physical Activity Questionnaire (IPAQ). \| **MEDAS-14** \| \|; **IPAQ** \| **Low adherence** \| **Moderate adherence** \| **High adherence**; **Low** \| Balance \| Exercise \| Exercise; **Moderate** \| Diet \| Balance \| Exercise; **High** \| Diet \| Diet \| Balance Each of the three possible profiles receive a different number of SMSs about diet and exercise attending this proportion: \| **Medication** \| **Diet** \| **Physical Activity** \| **Diabetes self-management**; **Balance** \| 50% \| 20% \| 20% \| 10%; **Diet** \| 50% \| 30% \| 10% \| 10%; **Exercise** \| 50% \| 10% \| 30% \| 10% Some messages include hyperlinks to different online resources of interest to people with type 2 diabetes. Both, text messages and linked online resources are in Spanish. An informative sample of DiabeText text messages are available upon request to the corresponding authors. |
| 4. | Procedures: Describe each of the procedures, activities, and/or processes used in the intervention, including any enabling or support activities. | 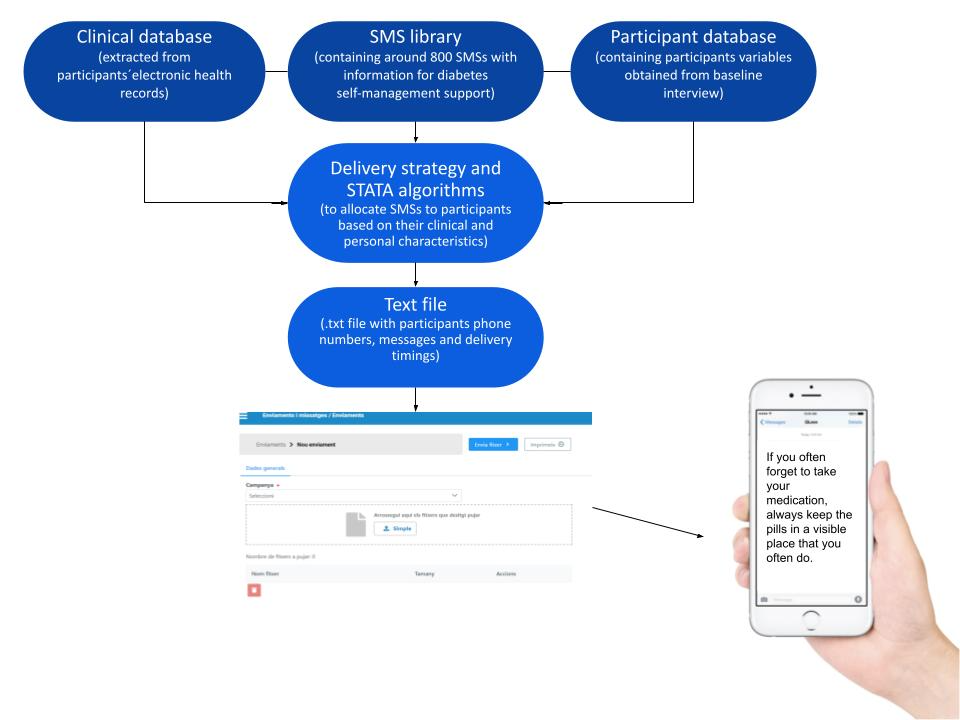  The DiabeText system is based on a set of algorithms that merge three input sources (clinical databases, database of brief messages, and patient-reported data) to generate text files (outputs) containing the text messages that each patient would receive on a certain day based on their clinical and behavioural data. The resulting .txt files are then manually uploaded to the SMS platform of the Balearic Islands Health Service which sends them to the patients. |
| 5. | **WHO PROVIDED**  For each category of intervention provider (e.g. psychologist, nursing assistant), describe their expertise, background and any specific training given. | The DiabeText intervention is designed to work autonomously after physicians’ prescription. In the research context, the enrolment of patients is assisted by the project staff. |
| 6. | **HOW**  Describe the modes of delivery (e.g. face-to-face or by some other mechanism, such as internet or telephone) of the intervention and whether it was provided individually or in a group. | Individual short text messages (160 characters maximum) sent to mobile phones of participants |
| 7. | **WHERE**  Describe the type(s) of location(s) where the intervention occurred, including any necessary infrastructure or relevant features. | DiabeText is a mobile health intervention without a specific location to be held. However, participants should remain in Spain during the study because the technological system is not able to send SMSs out of national borders without extra costs. The communication channel is unidirectional and asynchronous. |
| 8. | **WHEN and HOW MUCH**  Describe the number of times the intervention was delivered and over what period of time including the number of sessions, their schedule, and their duration, intensity or dose. | Each participant receives one daily SMS between Monday to Friday during the first four months. Then, 3 SMS per week the following 4 months and after that, the frequency diminishes to 2 SMS per week for the last 4 months of the intervention period. Participant also receive extra SMS when new information is registered in their electronic health records about upcoming appointments in primary care, next drug dispensing at the pharmacy, blood test results for A1C and body weight. |
| 9. | **TAILORING**  If the intervention was planned to be personalised, titrated or adapted, then describe what, why, when, and how. | The intervention is personalised based on data extracted from electronic health records (clinical data) and baseline interviews (behavioural data). First, based on patient-reported data during the baseline interview, participants are allocated to one of the three different profiles available (balance, diet, exercise), receiving a different number of SMSs about diet and exercise respectively -see point three above). Secondly, participants receive personalized SMSs according to the antidiabetic medication registered in their electronic health record. Third, the receive personalized SMSs depending on a list of variables of personalization (IMC, use of the internet on the mobile phone, diabetic foot, cholesterol higher than 200mg/dl, IPAQ score, smoking status, hypertension, chronic kidney disease, working status, Ramadan follow-up). Finally, the SMSs that had less than 130 characters include the name of the participant at the beginning. |
| 10. | **MODIFICATIONS**  If the intervention was modified during the course of the study, describe the changes (what, why, when, and how). | No modifications were introduced in the DiabeText intervention during the study |
| 11. | **HOW WELL**  Planned: If intervention adherence or fidelity was assessed, describe how and by whom, and if any strategies were used to maintain or improve fidelity, describe them. | Research staff checked that all participants are receiving the intervention correctly by using the *Bitmessage* platform (which keeps a record of the SMS successfully delivered). We also programmed the intervention to send messages to two phone numbers from the research team with anonymous data from two participants in the control group, to check that SMSs are correctly sent and received. At the end of follow-up, we asked all participants in the intervention group if they received the SMSs properly. |
| 12. | Actual: If intervention adherence or fidelity was assessed, describe the extent to which the intervention was delivered as planned. | According to the meta-data extracted from the Bitmessage message platform, 66,993 (98.4%) out of the 68,092 SMS uploaded to the platform immediately reached participants’ phones. 177 (0.26%) were put on hold (mainly due to poor mobile coverage) and delivered later, and 922 (1.35%) were not delivered (mainly because the operator could not send them or due to wrong number). Therefore, 318 out of 334 (95.2%) participants in the intervention group who completed the study received more than 90% of the intervention (>165 messages in one year). One participant received less than half of the intervention (received 62 messages in one year) and one who did not receive any message. |
| 13. | Other important information about the intervention | - Theoretical framework: Behavioral change wheel - Design process was based in the Medical Research Council guidance for development and evaluation of complex interventions and has been fully described in previously published work:  1. **Zamanillo-Campos R**, Fiol-DeRoque MA, Serrano-Ripoll MJ, Mira-Martínez S, Ricci Cabello I. Development and Evaluation of Diabetext, a Personalized Mhealth Intervention to Support Medication Adherence and Lifestyle Change Behaviour in Patients with Type 2 Diabetes in Spain: A Phase II Pragmatic Randomized Controlled Clinical Trial. *International Journal of Medical Informatics*. 2023. 2. **Zamanillo-Campos R**, Zaforteza-Dezcallar M, Boronat-Moreiro MA, Leiva-Rus A, Ripoll-Amengual J, Konieczna J, Fiol-DeRoque MA, Ricci Cabello I. Non-adherence to non-insulin glucose-lowering drugs: prevalence, predictors and impact on glycemic control and insulin initiation. A longitudinal cohort study in a large primary care database in Spain. *European Journal of General Practice*. 2023. 3. **Zamanillo-Campos R**, Serrano-Ripoll MJ, Taltavull-Aparicio JM, et al. Perspectives and Views of Primary Care Professionals Regarding DiabeText, a New mHealth Intervention to Support Adherence to Antidiabetic Medication in Spain: A Qualitative Study. *Int J Environ Res Public Health*. 2022;19(7):4237. 4. **Zamanillo-Campos R**, Serrano-Ripoll MJ, Taltavull-Aparicio JM, et al. Patients’ Views on the Design of DiabeText, a New mHealth Intervention to Improve Adherence to Oral Antidiabetes Medication in Spain: A Qualitative Study. *Int J Environ Res Public Health*. 2022;19(3):1902.   Developers and owners: The researchers Ignacio Ricci-Cabello, Rocío Zamanillo-Campos, Maria Jesús Serrano-Ripoll, Elena Gervilla-García and Maria Antonia Fiol-deRoque from the Health Research Institute of the Balearic Islands. |

EHRs; electronic health records, IPAQ; six-item International Physical Activity Questionnaire, MEDAS-14; 14-point Mediterranean diet adherence screener, SMSs; short text messages, A1c; glycated hemoglobin.
