## Supplementary material 2 for "Effectiveness of DiabeText, a mHealth intervention to support diabetes self-management: randomized controlled trial in primary care"

**Supplementary material 2.** Details on outcomes’ collection and calculations.

A. **DATA EXTRACTED FROM ELECTRONIC HEALTH RECORDS**

**Glycated hemoglobin (HbA1c)**

HbA1c was extracted as percentage which is calculated following the standard formulae [HbA1c(%)=(HbA1c(mmol/mol)+23.5)/ 10.93] based on its concentration in blood samples. It identifies average plasma glucose concentration (1).

The most recent HbA1c data registered between 6^th^ of April and 12^th^ of November in 2021 were extracted for all participants at baseline.

At post-intervention, we extracted the most recent data which was available between 21^st^ September 2022 and 21^st^ February 2023.

**Medication possession ratio (MPR)**

We calculated adherence in terms of medication possession ratio (MPR), defined as the number of days with treatment as medication being dispensed from the pharmacy to the patient (numerator), out of the total days of treatment prescribed by the doctor (denominator) (2):

MPR = [Days with treatment (prescription dispensed) / Days with treatment as prescribed by the doctor] x 100

At baseline it was calculated as the mean adherence for all the glucose lowering drugs prescribed during the 6 months previous to recruitment excluding insulin.

At post-intervention it was calculated as the mean adherence for all the glucose lowering drugs prescribed during the 12 months follow-up excluding insulin.

**Medication adherence based on MPR**

Adherence was stated as MPR≥80% and non-adherence was considered when MPR<80% (3,4).

B. **DATA COLLECTED FROM TELEPHONE INTERVIEWS**

**Self-reported adherence to oral glucose lowering drugs**

Self-reported adherence to glucose medications was measured with a 7-items ad hoc questionnaire adapted from Chaves-Torres et al. (5) for people with T2D. Participants who obtained 7 points were considered adherent while the ones with < 7 points were non-adherent.

**Health-related quality of life**

The 5-level EuroQol 5-dimensional questionnaire (EQ-5D-5L) questionnaire (6,7) was completed at baseline and post-intervention interviews. The index score was calculated using STATA syntax code and values with the Spanish value according to Ramos-Goñi JM et al. (8).

**Self-efficacy to manage diabetes**

The validated scale known as diabetes management self-efficacy scale in Spanish (DSES-S) was completed at baseline and post-intervention interviews. The score for each item was the number circled. If two consecutive numbers were circled, the lower number (less self-efficacy) was coded. If the numbers were not consecutive, the item was not scored. The score for the scale is the mean of the eight items. If more than two items were missing, we did not score the scale following instructions. Higher number indicates higher self-efficacy (9).

**Adherence to Mediterranean Diet**

The 14-point Mediterranean Diet Adherence Screener (MEDAS-14) (10) questionnaire was registered at baseline and post-intervention. Participants were classified as low adherents (≤5), moderate adherents (6 to 9 points) or high adherents (≥10 points).

**Physical Activity**

A 6-items adapted from the short version of the International Physical Activity Questionnaire (IPAQ) (11) was registered at baseline and post-intervention. Participants were classified as having a low, moderate or high level of physical activity based on metabolic equivalent of task (METs) calculation and the rules described below:

High

Any of the following 2 criteria:

• Vigorous physical activity on at least 3 days and accumulating at least 1500 MET-minutes/week, or

• 7 or more days of any combination of walking, moderate, or vigorous activities accumulating a minimum of 3000 MET-minutes/week.

Moderate

Any of the following 3 criteria:

• 3 or more days of vigorous activity of at least 20 minutes per day

• 5 or more days of moderate-intensity activity and/or walking at least 30 minutes per day, or

• 5 or more days of any combination of walking, moderate or vigorous activities achieving a minimum of at least 600 MET-minutes/week.

Low

• No reported activity

• Does not meet categories 2 or 3.

**Participants’ satisfaction with the intervention and potential related harms**

We asked the following questions to participants in the intervention group at 12 months follow-up:

1. Based on your experience, do you think that receiving informational text messages about diabetes on your mobile phone is a useful tool to help you improve your diabetes care? Scale from 1 (Not at all useful) to 10 (Very useful)
2. Have you found it easy to access the messages you received? Scale from 1 (Not easy) to 10 (Very easy).
3. Have you enjoyed receiving information through your mobile phone during this time? Scale from 1 (Not enjoyed at all) to 10 (Totally enjoyed).
4. Do you think that receiving messages to improve diabetes management for 1 year has caused you any harm? Yes/No. If the answer is affirmative, describe the harm experienced.
