## Supplementary material 3 for "Effectiveness of DiabeText, a mHealth intervention to support diabetes self-management: randomized controlled trial in primary care"

**Supplementary Material 3.** Results obtained in the analysis with data not imputed.

Table S2. Association between the DiabeText intervention and glycemic control, diabetes self-efficacy and quality of life.

|  | Baseline | | 12 months follow-up | | Association coefficient estimates for participants in the intervention group compared to controls | |
| --- | --- | --- | --- | --- | --- | --- |
|  | Control (n=371) | Intervention (n=371) | Control (n=340) | Intervention (n=334) | Beta (95%CI) | p-value^1^ |
| Glycemic control (Hba1c (%)), median (IR) | 8 (7.6-8.8) | 8.1 (7.7-8.7) | 7.4 (6.7-8.3)^2^ | 7.5 (6.7-8.2)^2^ | -0.044 (-0.226 to -0.137) | 0.629 |
| Diabetes self-efficacy scale (DSES), median (IR) | 6.9 (5.9-7.9) | 6.9 (5.8-8) | 8 (6.6-8.9) | 8.6 (7.4-9.3) | 0.601 (0.375 to 0.826) | **<0.001*** |
| Quality of life (EQindex), median (IR) | 0.93 (0.89-1) | 0.93 (0.88-1) | 0.97 (0.89-1) | 1 (0.92-1) | 0.017 (0.005 to 0.029) | **0.006*** |

Analyses were performed using linear mixed-effects models adjusted for baseline values.

HbA1c, glycated haemoglobin; IR, interquartile range; CI, 95% coefficient interval

^1^p values and 95%CI based on percentiles using non-parametric Bootstrap

^2^ post-intervention HbA1c available for 321 patients in the control group and 309 in the intervention group

*Significance stated at p<0.05

Table S3. Association between the DiabeText intervention and self-reported adherence to medication treatment, adherence to Mediterranean diet and adherence to physical activity.

|  | Baseline | | 12 months follow-up | | Association odds for participants in the intervention group compared to controls | |
| --- | --- | --- | --- | --- | --- | --- |
|  | Control (n=371) | Intervention (n=371) | Control (n=340) | Intervention (n=334) | OR (95% CI)^1^ | p-value^2^ |
| Self-reported adherence to antidiabetic medication, n (%) | | | | | | |
| Non-adherent | 177 (47.7%) | 166 (44.7%) | 143 (42.1%) | 114 (34.1%) | 1 |  |
| Adherent | 194 (52.3%) | 205 (55.3%) | 197 (57.9%) | 220 (65.9%) | 1.385 (1.00 to 1.90) | **0.046*** |
| Adherence to Mediterranean diet, n (%) | | | | | | |
| Non-adherent | 90 (24.3%) | 100 (27.0%) | 171 (50.3%) | 163 (48.8%) | 1 |  |
| Adherent | 281 (75.7%) | 271 (73.0%) | 169 (49.7%) | 171 (51.2%) | 1.08 (0.80 to 1.49) | 0.613 |
| Adherence to physical activity, n (%) | | | | | | |
| Non-adherent (low level) | 126 (34.0%) | 131 (35.3%) | 146 (43%) | 163 (48.8%) | 1 |  |
| Adherent (moderate or hight level) | 245 (66.0%) | 240 (64.7%) | 194 (57%) | 171 (51.2%) | 0.80 (0.58 to 1.12) | 0.199 |

Analyses were performed using logistic regression adjusted for baseline values.

^1^ data represents Odds Ratio (OR) with 95% confidence intervals in parentheses (CI)

^2^ p values and 95%CI based on percentiles using non-parametric Bootstrap

*Significance stated at p<0.05
